# Intra-breath oscillometry detects ventilation inhomogeneity in Cystic Fibrosis children

**DOI:** 10.1101/2023.11.02.23297971

**Authors:** Tamara L Blake, Paul D Robinson, Maddison T Deery, Claire E Wainwright, Zoltan Hantos, Peter D Sly

**Affiliations:** Children’s Health and Environment Program, Child Health Research Centre, University of Queensland, South Brisbane, Australia; Department of Respiratory and Sleep Medicine, Children’s Health Queensland, South Brisbane, Australia; Department of Anesthesiology and Intensive Therapy, Semmelweis University, Budapest, Hungary

## Abstract

**Background:** Ventilation inhomogeneity (VI) is measured in patients with Cystic Fibrosis (CF) by lung clearance index (LCI) using the Multiple Breath Washout (MBW). In young children, feasibility is often low within busy clinical settings. Intra-breath oscillometry (IB-OSC) measures respiratory system reactance (Xrs), which is determined by the same physiological principles. Given the high feasibility of IB-OSC in young children (>80%), we aimed to explore whether Xrs variables reflected VI as measured by LCI.

**Methods:** Paired measurements of IB-OSC (TremoFLO C-100, 10Hz signal) and Nitrogen (N_2_) MBW (Exhalyzer D) were performed in 97 children with CF. Abnormal LCI was defined as ≥7.1. IB-OSC Xrs variables at end-expiration (XeE), end-inspiration (XeI), volume-dependence of Xrs (ΔX) and ΔX corrected for tidal volume (ΔX/VT) were compared between those with normal and abnormal LCI. Receiver operator characteristic (ROC) analysis was performed to identify cut-off values to detect abnormal LCI.

**Results:** 115 acceptable paired IB-OSC and MBW measurements were achieved by 85 children (male 61%, median age 7.8yrs [25^th^-75^th^% 5.8-11.9]). 52 (45%) measurements of LCI were abnormal. All Xrs variables where significantly decreased (more negative) in children with abnormal LCI compared to normal (XeE -2.98v-0.81; XeI -2.10v-1.00; ΔX -0.25v0.12; ΔX/VT -0.54v0.22; all p<0.001). Cut-off values for ΔX and ΔX/VT provided the best sensitivity and specificity for detecting abnormal LCI.

**Conclusion:** These results suggest that novel IB-OSC variables can detect abnormal VI as measured by N_2_ MBW-LCI. This technique may allow us to detect and monitor early changes to lung function in busy CF clinics more easily.

## Manuscript

Development of cystic fibrosis (CF) lung disease typically starts in the small airways during the early years of life, often before clinical symptoms become evident [1–3]. Conventional lung function techniques, such as spirometry, are unable to detect these changes in patients with CF [4]. The lung clearance index (LCI), measured from a multiple breath washout (MBW), provides an accurate measure of the unevenness of ventilation distribution (or ventilation inhomogeneity, VI) and serves as a more sensitive indicator of early CF lung disease and primary outcome in clinical trials [5, 6]. Integrating this technique into the busy clinical environment detrimentally affects feasibility due to its time-consuming nature with very young patients [7, 8]. Intra-breath oscillometry (IB-OSC), a modified OSC technique, tracks changes in respiratory system resistance (Rrs) and reactance (Xrs) within tidal breathing. IB-OSC also offers high feasibility rate (>85%) in lung function naïve children with shorter testing time [9]. IB-OSC examines data at specific points in the breathing cycle, notably at points of zero flow – end-expiration (eE) and end-inspiration (eI) – to calculate volume-dependent changes between these points, such as ΔX (XeE-XeI). Given that measures of Xrs reflect volume-dependent changes in individual lung units, the same principle that determines ventilation distribution, and offers improved feasibility and shorter testing time of IB-OSC in younger children, we propose that this technique could serve as a valuable screening tool in busy CF clinics to identify children likely to have abnormal LCI. We hypothesize that measures of Xrs collected using IB-OSC, particularly ΔX, will be sensitive in detecting abnormal LCI in this patient group.

We conducted paired IB-OSC and MBW measurements on 97 children (aged 3.3-16.7 yrs), between June 2020 and October 2022 during routine CF clinic visits at the Queensland Children’s Hospital; the largest CF clinic in the southern hemisphere. Written informed consent was obtained from all parents/guardians or participants if >18 years of age. Ethics was approved by the Children’s Health Queensland HREC.

Participants were classified as ‘symptomatic’ if they reported increased respiratory symptoms (cough, sputum production, evidence of lower respiratory tract infection) within the previous two weeks; otherwise, they were classified as ‘clinically stable’. IB-OSC measurements were performed (tremoflo® C-100, Thorasys, Canada) according to ERS guidelines [10]. During data collection, 20-second data recordings were collected using a single 10Hz sinusoidal waveform. Conventional OSC measurements using a 5-37Hz pseudorandom signal were also collected. Outcome variables included R5, X5, AX, ReE, ReI, ΔR, XeE, XeI, ΔX, and ΔX adjusted for tidal volume (ΔX/VT). Participants then performed nitrogen (N_2_) MBW (Exhalyzer-D, EcoMedics AG, Switzerland) in accordance with current ATS/ERS consensus statement recommendations [11]. FRC and LCI were reported as the mean of at least two valid trials, following reanalysis using Spiroware 3.3.1 software to correct for O_2_/CO_2_ sensor cross-sensitivity errors [12]. Abnormal LCI results were defined as measurements ≥7.1 [13]. We compared IB-OSC variables between parctipants with normal and abnormal LCI results using Mann-Whitney U tests. Receiver operator characteristic (ROC) analysis was used to determine sensitivity, specificity, likelihood ratios, and cut-off values of IB-OSC variables for detecting VI, as defined by abnormal LCI.

Overall feasibility was 94/97 (97%) for IB-OSC and 85/97 (88%) for MBW, with technically acceptable results achieved by 85/97 (88%) of children on both techniques. Among the remaining participants, nine were unable to achieve technically acceptable MBW but successfully completed IB-OSC, and three were unsuccessful on both techniques. The final analysis consisted of 115 paired measurements; 59 children contributed one set of paired measurements, 23 children contributed two sets, 2 children contributed three sets and 1 contributed four sets. On average, complete IB-OSC test sessions were achieved in 13 minutes (range 10-21 mins) compared to 40 minutes (range 22-73 mins) for MBW. LCI ranged from 5.22 to 14.87, with 52 (45%) measurements categorised as abnormal. No differences in age, height, weight, or genotype were seen between participants with normal vs. abnormal LCI. Statistically significant differences (p<0.05) were observed between children with normal vs. abnormal LCI results for R5, X5 and AX measured using conventional OSC, and ReE, ReI, XeE, XeI, ΔX and ΔX/VT measured using IB-OSC (Table 1). Overall, children with abnormal LCI results had significantly decreased (more negative) reactance variables compared to children with normal LCI. While resistance variables were elevated in children with abnormal LCI results, these were at a reduced magnitude in comparison to reactance.

**Table 1:**
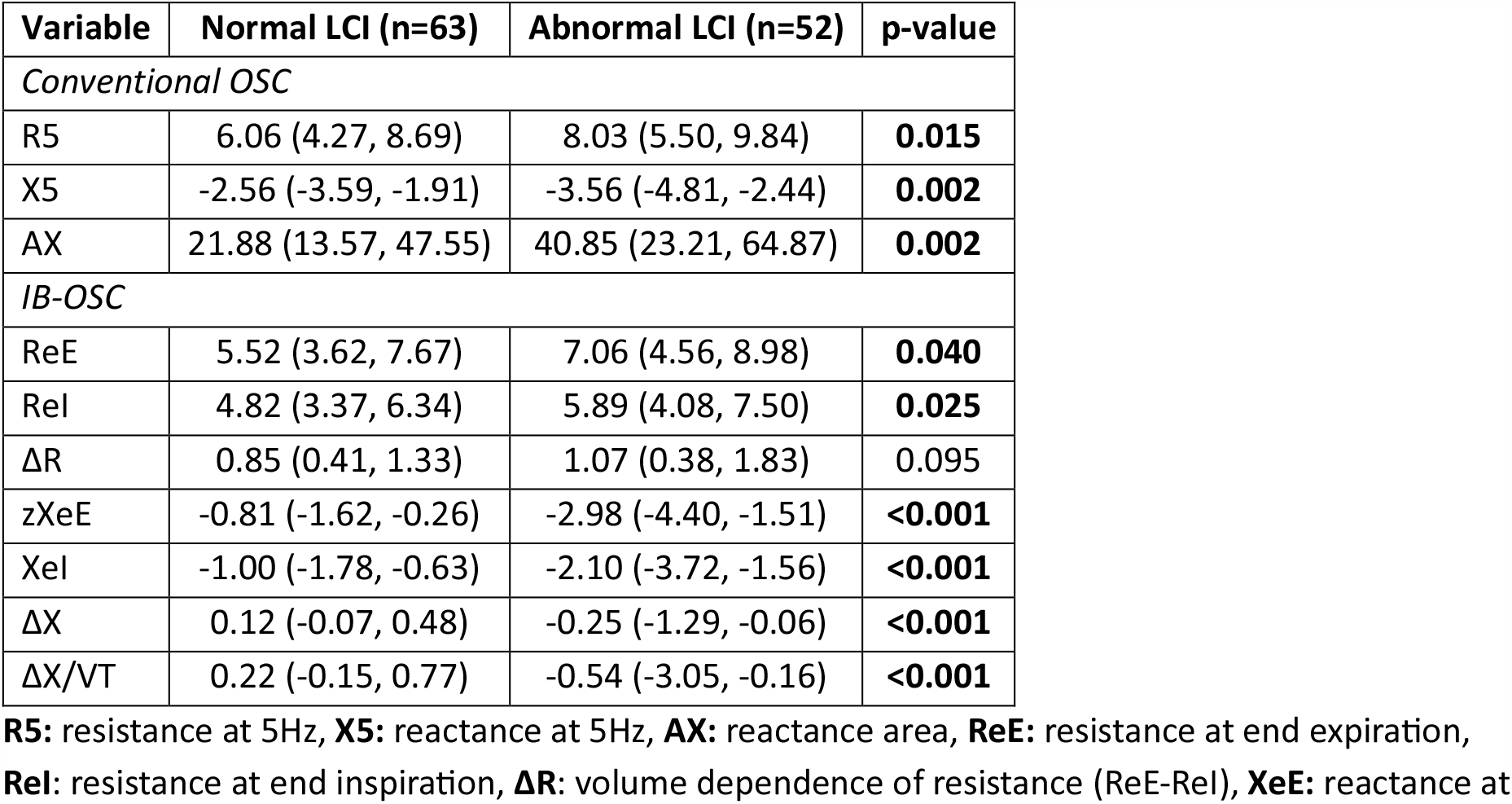
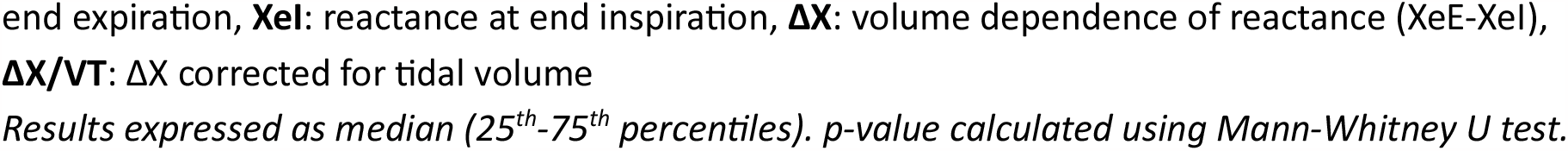
Comparison of convention and IB-OSC variables and normal vs. abnormal LCI results.

Figure 1 Panels A and B, display ROC curve analysis for all Rrs and Xrs variables (conventional and IB-OSC) to detect abnormal LCI result. All conventional OSC (R5, X5, AX) and IB-OSC Rrs (ReE, ReI, ΔR) variables performed lower than IB-OSC Xrs variables. The greatest area under the curve (AUC) values were seen for ΔX and ΔX/VT (0.77 for both) with a cut-off value of -0.08 and -0.15 cmH_2_0.s/L, respectively. For ΔX, positive and negative likelihood ratios were 3.29 and 0.35. Findings were similar for ΔX/VT, with positive and negative likelihood ratios of 3.46 and 0.30. Using these cut-off values, ΔX correctly classified 73% of participants with abnormal LCI (kappa = 0.51, moderate agreement), while ΔX/VT performed slightly beter at 77% (kappa = 0.55, moderate agreement).

**Figure 1. Panel A:**
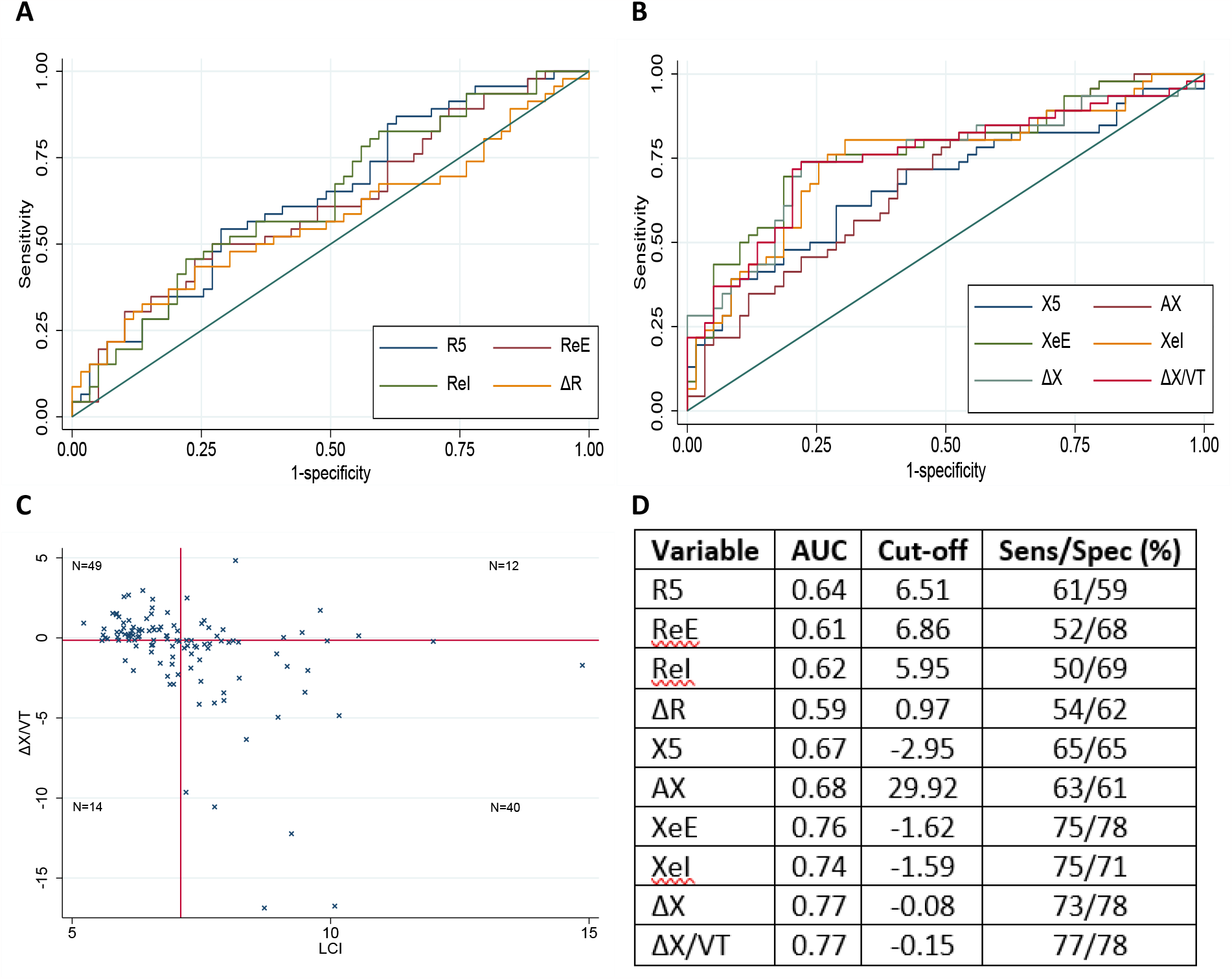
ROC curves for Rrs variables against LCI cut-off value (≥7.1). **Panel B:** ROC curves for Xrs variables against LCI cut-off value. **Panel C:** Scaterplot depicting ΔX/VT results against LCI for cohort. Vertical line denotes LCI cut-off value of ≥7.1. Horizontal line denotes cut-off value for ΔX/VT (−0.15 cmH_2_O.s/L^-2^) generated by ROC analysis. Overall concordance of ΔX and ΔX/VT was 75.6% and 77.4% respectively. **Panel D:** Table depicting AUC values, cut-off values and sensitivity/specificity scores for all variables.

When examined as an entire cohort (n=115), there was a 76% concordance between MBW and IB-OSC results using the identified cut-off points (generated from ROC analysis) for ΔX (43% classified as normal and 33% classified as abnormal in both tests) and a 77% concordance for ΔX/VT (Figure 1 Panel C, 43% classified as normal and 35% classified as abnormal in both tests). Similar trends were observed when examining clinical status (stable vs. symptomatic) at the time of testing. For clinically stable visits (n=68), there was a 78% concordance for ΔX (53% classified as normal and 25% as abnormal in both tests) and a 79% concordance for ΔX/VT (53% classified as normal and 27% as abnormal in both tests). For symptomatic visits (n=47), concordance was 72% for ΔX (28% classified as normal and 45% as abnormal in both tests) and 79% for ΔX/VT (28% classified as normal and 47% as abnormal in both tests).

Despite LCI correlating well with several clinically relevant outcomes [14], MBW remains challenging to successfully complete within busy clinical settings. The gold standard for MBW acceptability is three acceptable trials however, a recent study involving MBW-naïve young children found that only 24% could achieve three acceptable repeat measurements within 20 minutes [7]. Attempts to improve test feasibility have included reporting data from two valid trials rather than three [15, 16]. In our study, participants who were unable to achieve at least two MBW trials where all <7 years of age (success rate 87.6%). Of these, only three were also unable to achieve acceptable IB-OSC results (success rate 96.9%) and 96% of sessions were performed in less than 20 minutes.

This study represents the first direct comparison of IB-OSC Xrs variables and MBW LCI in the literature. MBW is the current gold standard for assessing VI in CF. All MBW and IB-OSC measurements were performed by highly trained technicians with significant experience in performing lung function testing in both preschool and school-aged children. Our cohort comprises approximately 40% of the total CF population at Queensland Children’s Hospital (the largest paediatric CF clinic in Australia) and was representative of the overall clinic with respect to demographics, disease severity, and management pathways (data not shown). We captured not only a wide age range but also a range of lung disease severities across mild to moderate impairment where MBW is currently used. Whilst we did not design this study to be a formal feasibility assessment, we provide clear evidence of the shorter test duration of IB-OSC, and report higher success rates, although this may be biased by the fact that IB-OSC was always performed first.

Our findings in this discovery cohort support our hypothesis that IB-OSC Xrs measures, particularly ΔX and ΔX/VT, are comparable to LCI results. Based on our preliminary data, ΔX and ΔX/VT values more negative than -0.08 and -0.15 cmH_2_0.s/L respectively, could be used to identify children likely to have elevated LCI results, thus facilitating targeted MBW testing in busy CF clinical settings. Future work will include confirming our findings in suitable validation cohorts.

## Data Availability

All data produced in the present study are available upon reasonable request to the authors.

